# Relationship between national changes in mobility due to non-pharmaceutical interventions and emergency department visits due to pediatric acute respiratory infections during the COVID-19 pandemic

**DOI:** 10.1101/2022.06.16.22276017

**Authors:** Franco Díaz, Cristóbal Carvajal, Sebastian Gatica, Pablo Vásquez-Hoyos, Roberto Jabornisky, Richard Von Moltke, Juan Camilo Jaramillo-Bustamante, Federico Pizarro, Pablo Cruces

**Author notes:** Corresponding authors: Pablo Cruces, MD. Camino A Rinconada 1201, Santiago, Chile. E-mail address, Franco Díaz, MD, MBA. Camino A Rinconada 1201, Santiago, Chile.

## Abstract

**Background:** Strong social distancing measures were quickly implemented in Chile during the SARS-CoV-2 outbreak. One of the aims of non-pharmaceutical interventions (NPI) mandates was to decrease overcrowding, thus is usually measured as mobility changes.

**Methods:** we gather data from national health statistics for pediatric emergency (PED) visits for acute respiratory infection (ARI) in children younger than 15. We defined a historical cohort, including data from 2016 to 2019, and compared them with 2020 and 2021 pandemic years. Also, Chile’s national mobility reports from the online google database were downloaded. We tested the correlation between changes in mobility and relative reduction in PED-ARI by Spearman’s Rank Test.

**Results:** Historical data showed a mean of 46863 ± 3071 PED-ARI weekly visits with a high seasonal variation, with two peaks in weeks 20 and 28 and weeks 32 to 36. This transient drop was temporally associated with the mid-winter 2-week holiday of schools. The usual PED visits peaks did not occur in 2020 and 2021. Mobility declined from week 9, reaching lower than historical data from week 12 and a minimum of 43% in week 15 of 2020 (Figure 1).

The correlation between mobility and PED-ARI visits showed a strong monotonic relationship (quadratic) with a Spearman’s rho of 0.80 (95% CI 0.75 to 0.86) (Figure 2).

**Conclusion:** NPI resulting in a decrease in mobility should be considered a robust public health measure to relieve the winter’s collapse of the national health system, decreasing morbimortality in children due to PED-ARI.

**WHAT’S KNOWN ON THIS SUBJECT:** A remarkable decrease in pediatric respiratory infections has been described during the pandemic, although the causes are still poorly understood.

**WHAT THIS STUDY ADDS:** In a historical cohort before the pandemic, we observed a temporal association between mid-winter holidays and the gap between the two peaks of acute pediatric respiratory infections. We found a strong correlation between national mobility changes due to non-pharmaceutical interventions and acute pediatric respiratory infections during the pandemic. Therefore, timely implementation of non-pharmaceutical interventions might be considered as a robust public health measure to attenuate the seasonal epidemic of non-COVID viral acute respiratory infections. With these data, we wonder if the time has come to implement non-pharmaceutical interventions to mitigate the stress, and frequently collapse, of national health systems due to the increase in pediatric acute respiratory infection, placing children as a priority, and provide the best care to this vulnerable population.

## Background

Strong social distancing measures were quickly established for containment as COVID-19 pandemic was imminent during the first trimester of 2020. Chile implemented strict non-pharmaceutical interventions (NPI) as early as March 2020 and maintained most of them during the following two years^1,2^. One of the aims of NPI was to decrease overcrowding of places of work, shopping, and study, among others, usually quantified as changes in mobility. Chile presents a dramatic increase in the pediatric emergency department (PED) visits during winter due to the seasonal epidemic of viral acute respiratory infections (ARI), stressing the healthcare system and causing serious morbidity and mortality in the pediatric population.^3, 4^ Several studies worldwide showed a decrease in ARI during the pandemic^5-9^. Although the mechanism is still poorly understood, it suggests that NPI might have a role in the incidence of non-COVID-19 viral infections. Therefore, we hypothesized a relationship between changes in mobility and the decrease in PED-ARI visits.

This study aimed to correlate the variation of PED visits for ARI in children and changes in mobility due to NPI during the first two years of the pandemic.

## Methods

This retrospective study was conducted using online public databases. For PED-ARI visits, we interrogated National Health Statistics Database of Chile^10^ with the following variables:

1. Patients younger than 15 years old
2. PED discharge diagnosis of ARI
3. Dates expressed as epidemiological weeks between 01.01.2016 and 12.31.2021.

We gathered Chile’s national mobility data from Google Community Mobility Reports^11^ from April 8^th,^ 2020 (the registry’s beginning) till December 31^st,^ 2021. This database describes the mobility change over time captured by mobile devices.

We defined the historical cohort as all PED-ARI consults from 2016 to 2019. Pandemic years’ data, 2020 and 2021, were compared to the mean of historical cohorts.

The PED-ARI variation was calculated as the difference between the weekly mean of the data recorded from 2016 to 2019 and the 2020 and 2021 cases per epidemiological week, expressed as a percentage. The analysis did not consider the 53rd epidemiological week for leap years.

The school year calendars of Chile were obtained from the Ministerio de Educación of Gobierno de Chile website^12^. The most important milestones and health actions of the pandemic during the research period were reviewed from Ministerio de Salud de Chile Monograph^1^.

After the database was constructed, we tested the correlation between changes in mobility and relative reduction in PED-ARI by Spearman’s Rank Test.

## Results

Historical data showed a mean of 46863 ± 3071 PED-ARI weekly visits with a high seasonal variation. There is a nadir between weeks 5 and 8 and a zenith between weeks 23 and 26, increasing weekly ED visits to 4.2 times. There are two high ED visit periods during the winter, between weeks 20 and 28 and between weeks 32 to 36, increasing 3.3 and 3.5 times the annual average. School mid-winter break was at weeks 29 and 30 during 2016-2019.

During 2020, in week 9, there was a decrease in PED-ARI visits, reaching significance at week 12 and a minimum at week 33 during 2020 with a 32.7-fold reduction compared to historical data. The usual peak of consultations during winter did not occur in 2020 and 2021. The decrease in PED-ARI ER visits remained significant between week 12 of the year 2020 and week 44 of the year 2021.

Mobility declined from week 9, reaching lower than historical data from week 12 and a minimum of 43% in week 15 of 2020 (Figure 1).

**Figure 1.**
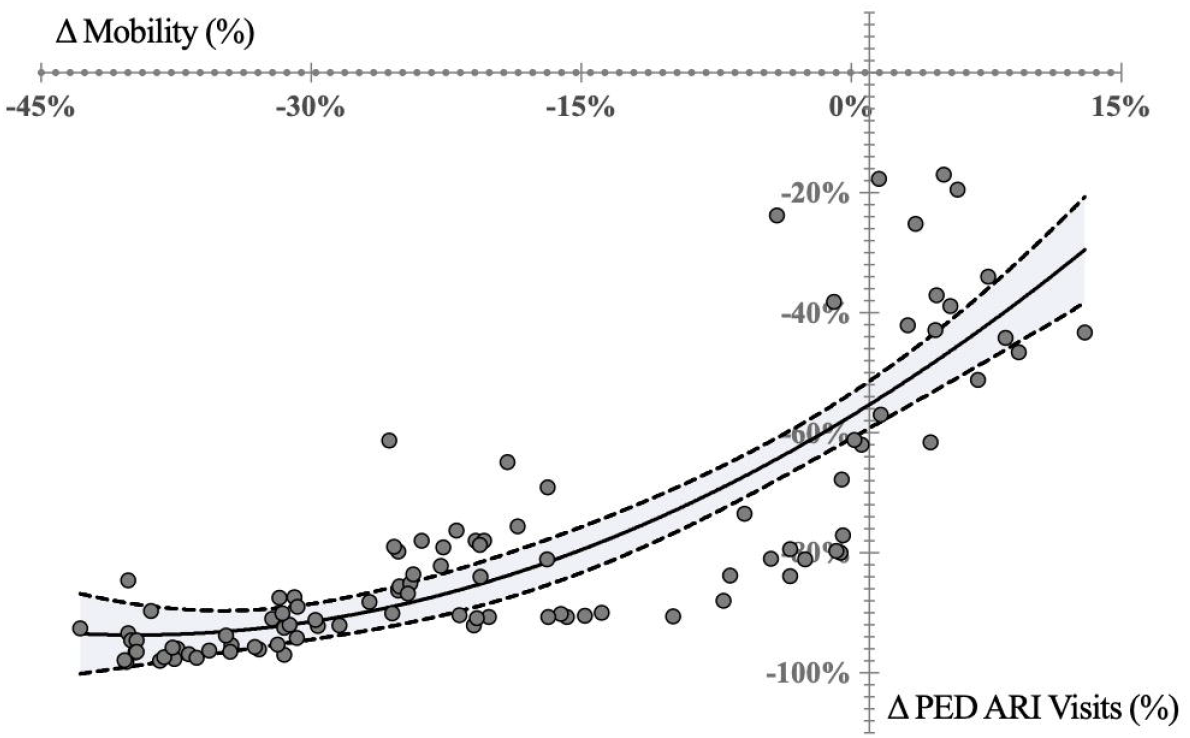
Variables analyzed per epidemiological week. (A) pediatric emergency visits for acute respiratory infections (PED-ARI) in Chile. Historical cohort, 2015-2019 average, and consecutive weeks of pandemic, 2020 and 2021. (B) Variation between historical cohort and consecutive weeks of the pandemic, 2020 and 2021, for PED-ARI; (C) National mobility variation per epidemiological week during 2020 and 2021. (0) First COVID-19 case in Chile; (1) Mandatory School closure (2) Partial quarantines in high incidence counties; (3) National Catastrophic Disaster declaration; (4) total quarantine in major urban areas; (5) DELTA variant identified in Chile; (6) Universal Vaccination for > 16 yo; (7) Vaccination for > 12 yo; (8) Non-mandatory in-person schooling; (9) Sanitary passport for vaccination proof in >17 yo; (10) Vaccination for children 12-17 yo; (11) Vaccination for children 6-11 yo; (12) Sanitary Passport for children (12-17 yo); (13) Omicron first case in Chile; (14) Vaccination for children 3-5 yo.

**Figure 2.**
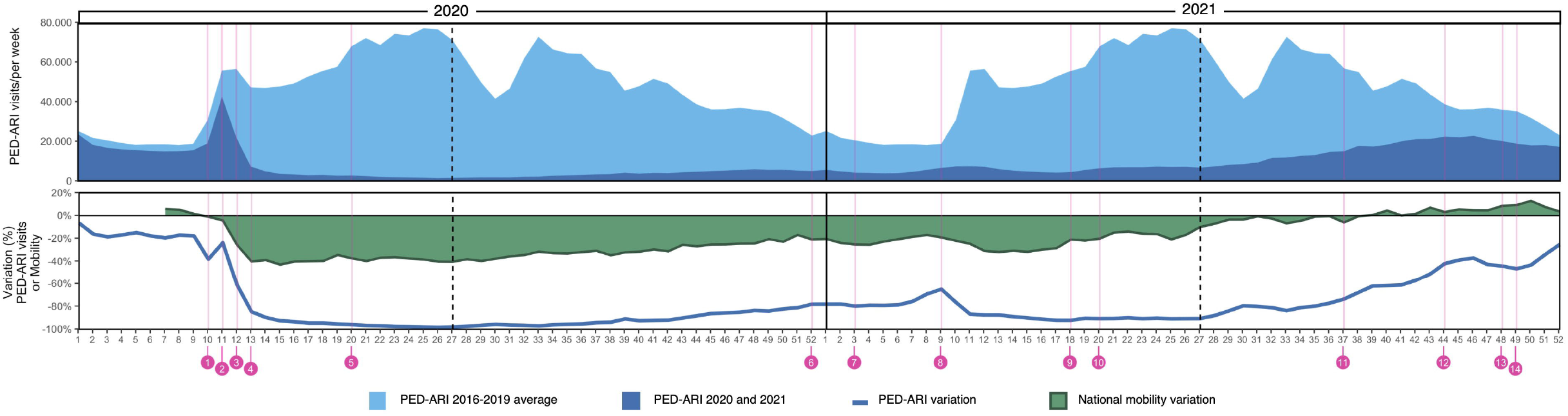
Correlation between national change in mobility (%) and the relative reduction in pediatric emergency department visits for acute respiratory infections (D PED-ARI). Spearman’s Rank Test, Spearman’s rho of 0.80 (95% CI 0.75 to 0.86).

## Discussion

Our study showed a correlation between mobility and PED-ARI during the first two years of the pandemic. When stricter NPI were implemented at the beginning of the pandemic, during southern hemisphere’s fall of 2020, there was a pronounced decrease in mobility, and then a blunt of the usual peaks of PED-ARI consults. NPI lost their effect as the pandemic progressed, and mobility and PED-ARI gradually returned to baseline.

The transient decrease of PED consults that historically results in a double-peak during winter is temporally associated with school mid-winter break of two weeks. In accordance with the historical data, the NPI during the pandemic resulting in a decrease in mobility, had a close relationship with the dramatic reduction in PED-ARI.

Respiratory admissions during the pandemic have been previously reported, but in our analysis, we were able to show a correlation with the net result of NPI that decreased mobility. Furthermore, the quadratic (non-linear) relationship highlights that the initial mobility change has a more powerful effect on reducing PEDA-ARI ED visits and then loses its effectiveness.

This study’s implications go beyond the COVID-19 pandemic. Every year the increase of PED-ARI frequently causes the national health system’s collapse. Surprisingly, despite having robust data on this issue for more than 25 years, the measures are focused on reinforcing the capacity to respond to the increase in demand^4^. Although, any action becomes inefficient and ineffective, facing an increase of more than four times the demand over a couple of weeks.

NPI might be a robust public health measure to attenuate the massive increase in PED-ARI consults and admissions during critical winter epidemiological weeks. ARIs are the third cause of death and the first cause of admissions in children younger than five years in Chile and one of the leading causes of morbimortality in infants worldwide ^13-16^. Preventive measures might be crucial in PED-ARI because access to proper care in many places in Latin America and the world is non-available or difficult. Even more, in non-developed regions, there is a correlation between PED-ARI mortality and socioeconomic status between and within countries^17-21^. Recent data show that admission due to PED-ARI has a negative impact on development, even in high-income countries. ^22^

Our study has some limitations. Many factors contribute to the incidence and the winter’s peak of ARI, like the climate characteristics, the socioeconomic status, and the prevalence of chronic diseases in the population. Although, it is reasonable to think that most of them did not significantly change before and after the COVID-19 pandemic. This study is a secondary analysis of previously collected data, thus, accuracy and bias cannot be evaluated. Given the retrospective analysis causative effect or its magnitude cannot be concluded.

Despite these limitations, we demonstrated how mobility restriction, one of the NPIs adopted during the pandemic, could be responsible for decreasing PED-ARI. Furthermore, the decrease in mobility had a quadratic correlation, so the initial restrictions had a more significant effect on PED-ARI than extreme restrains, like total quarantine.

The first two years of the pandemic proved to be a large-scale social and health laboratory, implementing empirical *gerontocentric* measures^23^. Despite the devastating effects on children and adolescents, it could lay the groundwork for future innovative NPI during the seasonal epidemic of viral acute respiratory infections. With these data, we wonder if the time has come to timely implement NPI to mitigate the winter’s stress and collapse of the healthcare systems due to the massive increase in PED-ARI, prioritizing children, and providing the best care to this vulnerable population.

## Data Availability

Data for this study was gathered exclusively from public databases:
- Departamento de Estadisticas e Informacion de Salud, DEIS. Ministerio de Salud, Gobierno de Chile. Available at < https://deis.minsal.cl/#estadisticas >
- Google Community Mobility Reports, Available at:
< https://www.google.com/covid19/mobility/ >

https://www.google.com/covid19/mobility/

https://deis.minsal.cl/#estadisticas

## Ethics approval

Data for this study was gathered exclusively from public databases:

- Departamento de Estadísticas e Información de Salud, DEIS. Ministerio de Salud, Gobierno de Chile. Available at < https://deis.minsal.cl/#estadisticas >
- Google Community Mobility Reports, Available at < https://www.google.com/covid19/mobility/ >

## Data availability

Data for this study was gathered exclusively from public databases:

- Departamento de Estadísticas e Información de Salud, DEIS. Ministerio de Salud, Gobierno de Chile. Available at < https://deis.minsal.cl/#estadisticas >
- Google Community Mobility Reports, Available at: < https://www.google.com/covid19/mobility/ >

## Author contribution

FD proposed the original idea for the study, designed the study, supervised data gathering and analysis and drafted first and subsequent revised manuscripts.

and manuscript draft

CC made a substantial contribution to the design of the study, gathered, and analyzed the data, and participated in writing the results and manuscript draft.

SG made a substantial contribution to the analysis of data and manuscript writing and revision.

PVH performed the analysis of data, made a substantial contribution to the manuscript writing and subsequent revisions.

RJ made a substantial contribution to manuscript writing and subsequent editions and revisions

RVM made a substantial contribution to the analysis of data and manuscript writing and revision.

JCJB made a substantial contribution to manuscript writing and subsequent editions and revisions.

FP made a substantial contribution to the analysis of data and manuscript writing.

PC made a substantial contribution to the design of the study, participated in manuscript writing and subsequent editions and revisions.

All authors approved the final version of the manuscript

